# PREDICTIONS FOR EUROPE FOR THE COVID-19 PANDEMIC AFTER LOCKDOWN WAS LIFTED USING AN SIR MODEL

**DOI:** 10.1101/2020.10.03.20206359

**Authors:** Jay Patwardhan

## Abstract

I analyze a simplified SIR model developed from a paper written by Gyan Bhanot and Charles de Lisi in May of 2020 to find the successes and limitations of their predictions. In particular, I study the predicted cases and deaths fitted to data from March and its potential application to data in September. The data is observed to fit the model as predicted until around 150 days after December 31, 2019, after which many countries lift their lockdowns and begin to reopen. A plateau in cases followed by an increase approximately 1.5 months after is also observed. In terms of deaths, the data fits the shape of the model, but the model mostly underestimates the death toll after around 160 days. An analysis of the residuals is provided to locate the precise date of the departure of each country from its accepted data estimates and test each data point to its predicted value using a Z-test to determine whether each observation can fit the given model. The observed behavior is matched to policy measures taken in each country to attach an explanation to these observations. I notice that an international reopening results in a sharp increase in cases, and aim to plot this new growth in cases and predict when the pandemic will end for each country.

## 1. Introduction

The novel Sars-CoV-2 coronavirus, first appearing in Wuhan, China at around December 31, 2019, has caused an ongoing worldwide pandemic. It differs from the initial Sars-CoV virus strain in a multitude of ways, from its only 79 percent similarity to the original [8], to a wider range of mortality rates. While the Sars-CoV strain had about a 9.6 percent mortality[6], this new coronavirus has death rates ranging from 13.9 percent in Italy to 3.1 percent in the US[5]. Despite this, it appears to have a more favorable transmission rate and a prolonged latency period. Since the onset of a global lockdown earlier in the year, many countries throughout the world have begun reopening, albeit not in full. Nonetheless, the measures taken by governments during quarantine have significantly eased, although many countries continue to encourage proper social distancing measures, from wearing masks to staying 6 feet apart. Around 140 days after December 31, 2019, an SIR model[10] was created to model the flow of this novel coronavirus in the European countries of the Netherlands, Denmark, Sweden, Norway, the UK, Spain, Germany, Italy, and France[4]. The lack of a common policy amongst the world has led to different responses and outbreaks in different countries, as we will come to see. Varying reopening policies between each country has resulted in varying levels of success in virus containment, with some countries almost entirely shutting down the virus and other countries seeing a plateauing and subsequent growth in cases.

## 2. A summary of the Model

In my study, I focus on the two differential equations modelling the S and I part of the SIR model, meaning Susceptible and Infected, respectively. This uses the variables X1, denoting S, X2, denoting I, α, the transmission rate (number of infections per day per contact), *γ*, the rate at which individuals leave the infected population, N, the total individuals in an interacting pool of Susceptibles, δ, the fraction of individuals who die after being infected, and P, the maximum value of X2. The fits for the amount of deaths are found by scaling the cases by δ and shifting the graph forward by a certain amount of days.

## 3. Relevant Equations

All of the relevant equations used to build the model are taken from the paper that develops the model[4]. The primary equations used to graph the curves are:

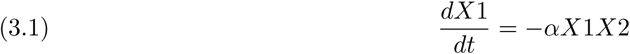

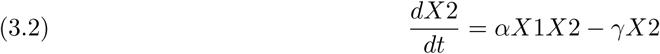

Finding the residuals themselves is an easy enough equation: If R = the value of the residual, x = the observed number of cases or deaths at a point in time, and X2 = the predicted number of cases or deaths at a point in time using the main curve, then

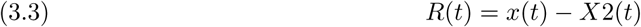

The associated error bars to each residual are found from subtracting the observed number of cases or deaths from the predicted number of cases or deaths at a point in time using each value of the error bar curve.

To each point in time, I also associate a p value denoting the probability of finding the observed data result if the model was true. This is a matter of a simple hypothesis test.

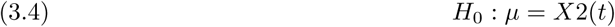

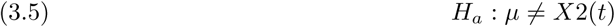

The standard deviation is found by using its formula and the values X2U and X2L representing the upper and lower bounds from the estimation model:

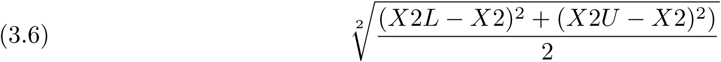

Using the formula, we can then find the corresponding z score and the area under the curve representing the p value using

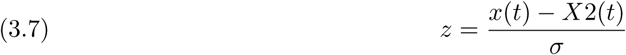

Alternatively, using the pnorm function in R yields the same results.

## 4. Results and Discussion

### 4.1. Reasons for Data Analysis tools

In this exploration, both residual plots and p value plots (at the α = 0.05 level corresponding to a 95 percent level of confidence) are utilized to find results. We can observe both the advantages and drawbacks to each tool, which provide a need for the other. With a residual plot, we can better analyze how far each data point falls from the predicted value on a linear scale. This clears up the confusion of distances on the log scale, as the model may show an overprediction of 1000 to be miniscule in comparison to an overprediction of 10 based on the data point’s location on the y axis. We also gain a better understanding of both how well the data fits the model and when the data starts and stops fitting the model. The drawback, however, is a lack of knowledge of whether a certain residual is expected from the data for the same reason that there is a distinction between the aforementioned difference of 1000 and 10. The error bars expand as time goes on, encompassing a larger expanse of area, but deceptively accounting for a less inclusive interval of error. It is useful to gather whether an observed data point’s deviation from the model is expected or not. A statistical analysis into the probability of observing such an extreme or more extreme data point tells us that even a large difference of 1000 can be expected given a large standard deviation as dictated by the error bars. A problem, however, arises when the R value derived from the model[4] increases. As R increases, the fit becomes tighter and tighter, decreasing the standard deviation, potentially making even a deviation of 10 seem probabilistically impossible. This is where a look at the residuals clears up the problems caused by small standards of deviation. In other words, residuals are particularly useful when studying smaller values, while p values are more useful when studying larger values. A final useful note is that all labeled days represent days starting after December 31, 2019 (January 1, 2020 is Day 1).

### 4.2. Analysis of the Case Model

In general, the model matches the observed data points well until around Day 150. We note an initial almost linear climb in log space, accounting for an exponential growth of the virus. We note a maximum point at a median of about 40 days after the first case detected in the country, followed by a more gradual drop in cases. Instead of seeing a continuous decrease in cases, we observe a plateau, sometimes followed by a small drop in cases, and then a rise which on occasion exceeds the initial peak. This plateau starts at different locations, most after an initial drop in cases, but some at the peak itself. The peaks of each country vary, and we can see a clear relationship between a country’s population and their number of cases. Looking at the number of cases per million population, however, does not show any such relationship, suggesting that this factor depends more on federal action rather than just population and population density. We notice that both the UK and Sweden plateau immediately after peaking, making the model mostly inaccurate for both countries. For this reason, we can’t do a p value analysis. Along a similar vein, the observed drop in cases for Italy didn’t match the model as the drop was much more gradual than expected, and for that reason the model is mostly inaccurate for this country as well. For every other country, the data points began to deviate from the model at about Day 142 *±* 20 days. These plateaus lasted for about 42.75 *±* 15 days, with the exception of Norway, with an 83 day plateau. Given the relatively small size of the outbreak in Norway, it is expected that there would be a thickly shaped plateau in cases due to the smallness of the numbers involved. It can also be argued that France doesn’t have a plateau at all, instead going from a decrease in cases to a sudden increase. Another interpretation would be to observe a wide plateau akin to the shapes of Norway and Denmark, followed by a rise in cases. We notice an increase in cases in every country, with most increases being sharply exponential (with the exception of Sweden). We also observe a small decline in cases right prior to the aforementioned increase (with the exception of Spain and Germany), suggesting that many countries had the potential to drop their cases once again before whatever event caused them all to grow so suddenly. Finally, we observe this rise in cases to occur roughly in the same timeframe: from Day 190 to day 205. We can likely attribute this to a policy either made by the EU or by each government in close proximity to the other regarding reopening. We’ll explore this in a later section. The residual plots were useful when attaching a rough start date to the plateau, as it roughly corresponds to a deviation from the Covid model. A point and its error bars not intersecting with 0 whose future points also do not intersect with 0 is thus marked as a starting point to locate these plateaus. As time went on, the model predicted the virus to be gone in most countries before Day 180[4], with the exception being the UK at around Day 210. After around Day 170, the model predicts small X2 values with small standards of deviation, so observed residuals at this point mostly represent the growth of the virus itself during that time. We can clearly see an exponential increase both in the model and in the residual plots at nearly identical times for this reason. Similarly, we mark the date in which the observed data points are first statistically improbable given the model curve and continue to be (mostly) improbable as the date at which point the model becomes obsolete using our p value plots. This also matches quite well with the start date of the plateau.

### 4.3. Analysis of the Death Model

We can observe a similar shape for each death graph as that of the cases graph, but with the values on the y axis scaled down by whatever fraction *δ* was found. We notice almost consistently that the model underestimates the total death rate after its peak, with the exception of the Netherlands, which the model surprisingly overestimates. The model accounts for deaths to be 0 at around 160 days for each country, but instead we see that there are still deaths after the predicted end date, albeit very few. We can see this in our residual plots, in which the graphs stabilize around 0 after around Day 160. This stabilization, however, is not an indicator of the accuracy of the model, which is proven by the p values dropping below the *α* = 0.05 level after roughly the same date, but because the deaths have also fallen to fairly small numbers, usually around 1-10. The model fails around 15 days after cases plateau, because we expect cases and deaths to decrease continually until they reach 0, but instead we get a stagnation of the virus. It wouldn’t be apt in most cases to call this persistence of the virus a plateau, but rather the failure of the model to graph a much less steep falling curve. We notice that the UK and Italy almost immediately deviate from the expected curve after reaching its peak, and we can see a much more gradual decrease in cases in both countries. In almost every residual plot, we see a shape resembling a damped harmonic oscillation curve as the residuals approach 0. As x approaches 200 and beyond, we can say that if the residuals don’t go to and stay at 0, there’s an anomaly. We observe this in the UK and in Sweden. In the UK, we notice a fluctuation between 0 and 125 from Days 163 onwards, indicating a stagnation in deaths. Each stagnation occurs during a period of increased cases. While we may expect an increase in cases to signify an increase in deaths, it might be useful to consider the possibility that the lethality of the virus itself has decreased, or hospitals have adapted and become better equipped to help patients survive the virus. We know that the virus takes a long time to mutate[9], and that its mutations are mostly inconsequential, so we could choose to focus our analysis on hospitals. During the peak of the virus, hospitals were mostly swabbing and testing severely symptomatic patients, arguably skewing the lethality of the virus. Considering that case rises could be associated with increased numbers of both moderately and severely symptomatic patients instead of just mostly severely symptomatic patients, we can infer that the number of deaths wouldn’t have increased[3]. In addition to this, younger people are getting the virus, leading to a greater survival rate, and hospitals themselves have become better at preventing deaths, due to the efforts of medical researchers. All of this has continued to keep deaths down. In fact, since we don’t see a surge in deaths as we see a surge in cases, it could be predicted that deaths would increase after day 227 in order to compensate for the rise in cases, but it is also entirely probable that the persistence and “plateau” that we see prior to day 227 is due to the surge of cases in each country. It is also probable that it takes longer to die now with the virus than it did before. Finally, an examination of the date of the first deaths in each country compared to the days until the model began to deviate from its expected behavior yields no relation, indicating that any failure of the observed data points to conform to the model is most likely due to the policy measures taken in each country.

### 4.4. Policy Measures taken in each Country

We have seen almost uniform behavior amongst all of the countries in the shape of their growth, but the days for each significant change in shape vary. This begs the question, what changed? We know that it takes about 15.5 days for a person who gets infected to be recognized as infected and to get removed from the population, so we can infer that any action taken will have repercussions about 15 days later. However, seeing as this number was recorded when mostly severely affected people were being diagnosed, it can be assumed that it’ll take less time in the months of July and August to get diagnosed. After about 183 days since December 31 2019 (July 1, 2020), the EU began lifting restrictions on nonessential travel for some countries. This has caused a direct increase in the number of cases for Italy, Germany, Spain, Norway, Denmark, the Netherlands, France, and to a much lesser extent, Sweden. We can see these effects in the rise in cases in each country from Days 190 to 205. The UK announced around 190 days since December 31 2019 that restaurants, bars, hotels, hairdressers, cinemas and museums would reopen, causing a surge in cases almost immediately. Sweden, which imposed a more voluntary lockdown, has experienced a more protracted outbreak than other countries, which we can see by the plateauing of cases in Sweden, with the exception of a sudden spike and drop in cases over the span of just a few days. In addition to this, every country on the list launched some phase of reopening in July and August, further compounding the issue caused by opening the borders[7]. This still begs the question, why did cases rise in this scenario but plateaued in prior days? This is likely because countries initially began lifting lockdowns internally during the Day 140-Day 160 period, launching their initial phases of reopening. These reopenings were largely internal, allowing for mobility inside the country. In Germany, smaller shops began to reopen, and the travel ban was lifted for EU member states. Italy ended travel restrictions, bars, restaurants, shops as well as other public activities such as tourist locations opened. Such changes happened to essentially every country, with restrictions being lifted in mid May (Day 130-Day 140). A smaller scale reopening coupled with falling cases likely contributed to a smaller growth of the virus causing a plateau, whereas larger scale reopenings in later months almost uniformly caused a significant rise in cases. Spain in particular has been hit hard, with many cases in Aragon[2], the most heavy hit county, due to infections with wandering seasonal fruit pickers. Nonetheless, while cases continue to increase, only about 3 percent of cases need hospital treatment, and the mortality rate is as low as 0.3 percent in some areas. This provides a cause for hope, but also indicates an inaccuracy in the Covid model in both its measurements of the fraction of cases that result in deaths and in the time it takes for an infected person to die.

### 4.5. A new model to chart the post Lockdown Case Growth

We then sought to chart the new growth of Covid-19 using the same equations as before. Using the times indicated by our analysis, we charted the new wave of cases. We found that the time before a person removes him/herself from the population (1/*γ*) increases dramatically, as well as the infectivity rate *α*. However, we notice no upward trends in deaths as of yet, and any increase in deaths is slow and not profound as of yet, suggesting that while cases are indeed on the rise, the death rate has stabilized. This can be linked to younger people getting the virus, as older age patients are more susceptible to receiving the virus[1]. An appropriate residual graph analysis shows that the residual cases have mostly dropped, with some exceptions. We can’t do a p value analysis due to the tightness of the graphs in addition to the flatter shape of the incline and the odd rises and falls due to the peculiarities of each country’s reopening format. We plot the best case scenario for each of these countries, as we see a general easing of the growth and a parabolic curve in each plot. We were unable to do so for Sweden due because we failed to observe a sharp rise in cases akin to the initial growth. The assumption thus is that the trend will continue downwards as it is predicted, which will likely require a continuation of the current reopening phase without the introduction of more radical changes that may shift the data. Studying the parameters, we observe that not only has the value for gamma significantly decreased for all of the countries, the value for alpha has too. In the Netherlands, the value for gamma drops from 0.05676 to 0.02647, in Denmark from 0.06 to 0.028, in Norway from 0.08 to 0.0355, in the UK from 0.0268 to 0.00716, in Germany from 0.0676 to 0.023, in Italy from 0.0556 to 0.0222, in Canada from 0.0267 to 0.008, in France from 0.06657 to 0.02, in the USA from 0.1676 to 0.00972, and in Spain from 0.689 to 0.0178. Similarly, in the Netherlands, the value for alpha drops from 9.07e-5 to 2.539e-5, in Denmark from 0.000329 to 6.205e-5, in Norway from 0.0005448 to 0.0003996, in the UK from 3.62e-5 to 8.528e-6, in Germany from 2.308e-5 to 2.472e-5 (increasing slightly in this scenario), in Italy from 2.588e-5 to 3.45e-5 (also increasing slightly here), in Canada from 7.94e-5 to 2.858e-5, in France from 2.889e-5 to 3.327e-6, in the USA from 1.04e-5 to 2.518e-6, and in Spain from 2.348e-5 to 4.141e-6. A decrease in gamma indicates that each infected person is taking more time to remove himself/herself from the population, suggesting that the symptoms and harm to them is significantly less. A lower alpha value indicates a lesser transmission rate, which aligns with the prediction that younger people are getting the virus. Younger people have a lesser history with preexisting conditions and are generally stronger and in better shape to fight the virus. If younger people are stronger in general and show less symptoms, it follows naturally that they would take longer to go to the hospital. Taking X2 < 5 to be the day that the pandemic ends, we conclude that it will end in the Netherlands on day 549 (July 2, 2021), in Denmark on day 505 (May 19, 2021), in Norway on day 383 (January 17, 2021), in the UK on day 804 (March 14, 2022 if we take 50 as our parameter), in Germany on day 384 (January 18, 2021), in Italy on day 580 (August 2, 2021), in Canada on day 712 (December 12, 2021 if we take 50 as our parameter), in France on day 750 (January 19, 2022), in the USA on day 900 (June 18, 2022 if we take 100 as our parameter), and in Spain on day 774 (February 12, 2022). While this is the expected fit, we hope for a vaccine to be found, or the development of herd immunity, which may reduce the time for the virus to be removed from the population.

## 5. Conclusion

This paper thus aims to provide a general shape to the future of the Covid-19 virus, and an examination of which policy measures resulted in a growth in cases. In almost every European country, we see the case model begin to fail at around Day 150, at which point the cases stagnate, then increase starting at around Day 190. This increase in cases is nowhere close to reaching the peaks of the initial outbreak, even though we see a sudden increase in the case count. This owes to the deceptive nature of space in the log scale, where a sudden increase from 400 to 900 is seen as more dramatic than an increase in the thousands. Spain, however, is dealing with another major outbreak, with cases approaching that of its first peak. The model also largely underestimates deaths and the continued persistence of deaths after Day 160. It also overestimates the lethality and underestimates the time to die in later months after around June. The deviation of the observed data points from the expectations set by the model of the model can be understood to be due to its inability to account for reopening, instead assuming that each country would remain in a state of total lockdown until deaths go to 0. It also fails to account for the outbreak amongst younger people, and thus overestimates the survival rate. Nonetheless, before the lifting of lockdown, the model fits the data quite well. An attempt to extrapolate the results of this model to Asian countries was attempted, but many countries there are still fighting the virus, seeing a growth in cases. Others observed a small peak around April, and are suddenly seeing a dramatic resurgence in August, akin to the peaks of the European countries in April. While the model could be adapted to the cases in August, an analysis of the data wouldn’t be appropriate until much after cases have subsided. Nonetheless, this paper aims to change the notions of the general public as to the situation of the Covid-19 virus, that contrary to popular opinion, it is still at large, and affecting a younger crowd. It also aims to provide an answer to what forms of reopening are still acceptable to reducing net cases, to which mostly internal reopenings are suggested. It finally looks to model the new growth of Covid-19 in the 9 European countries in addition to two new North American countries.

## Declaration regarding data and software

The data used in this paper were all derived from public sources. Links to these data are included in the paper. The main data source is taken from https://ourworldindata.org/coronavirus-source-data. The R code used to analyze the data along with all data files will be provided on request -.

**Figure 1:**
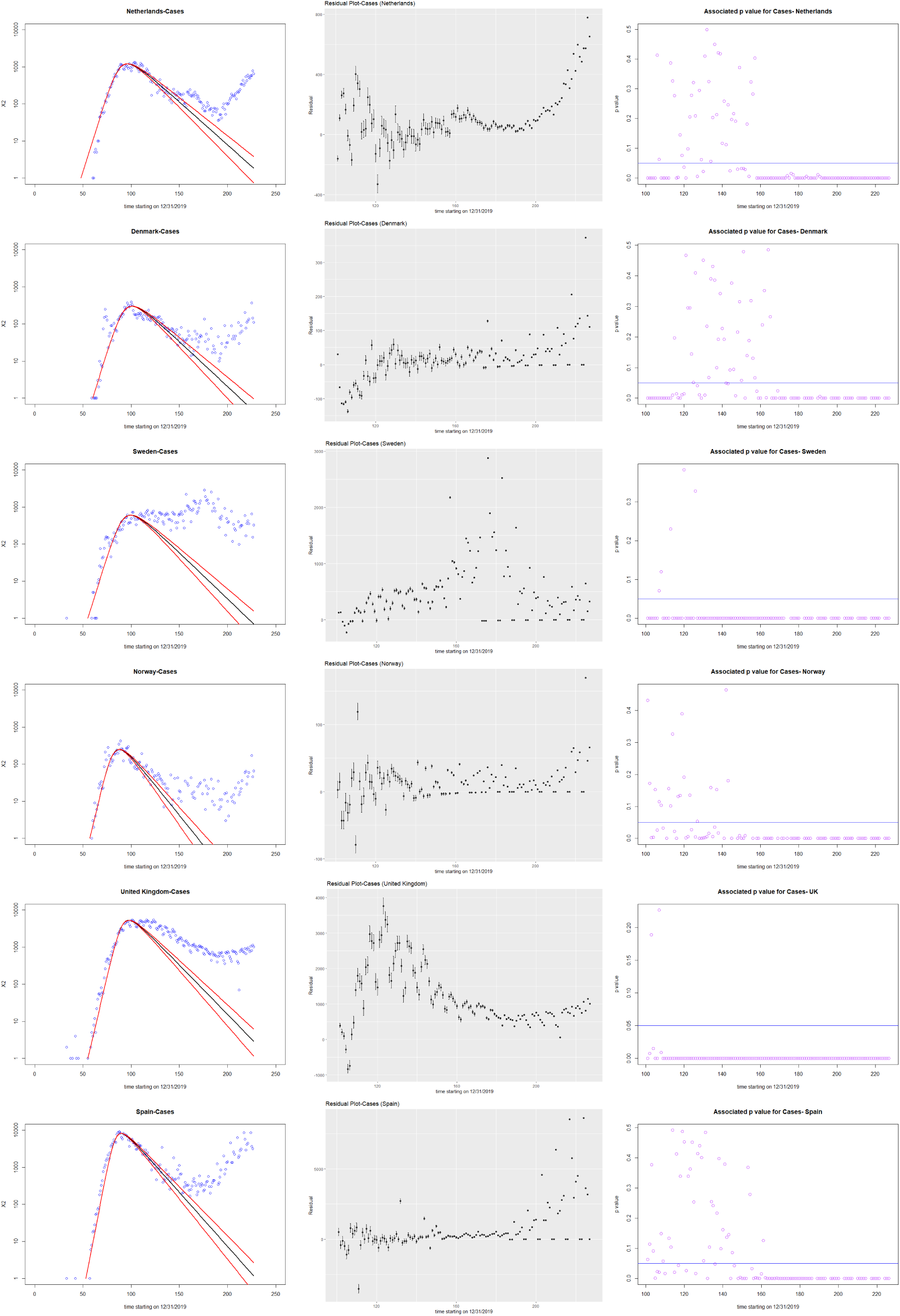
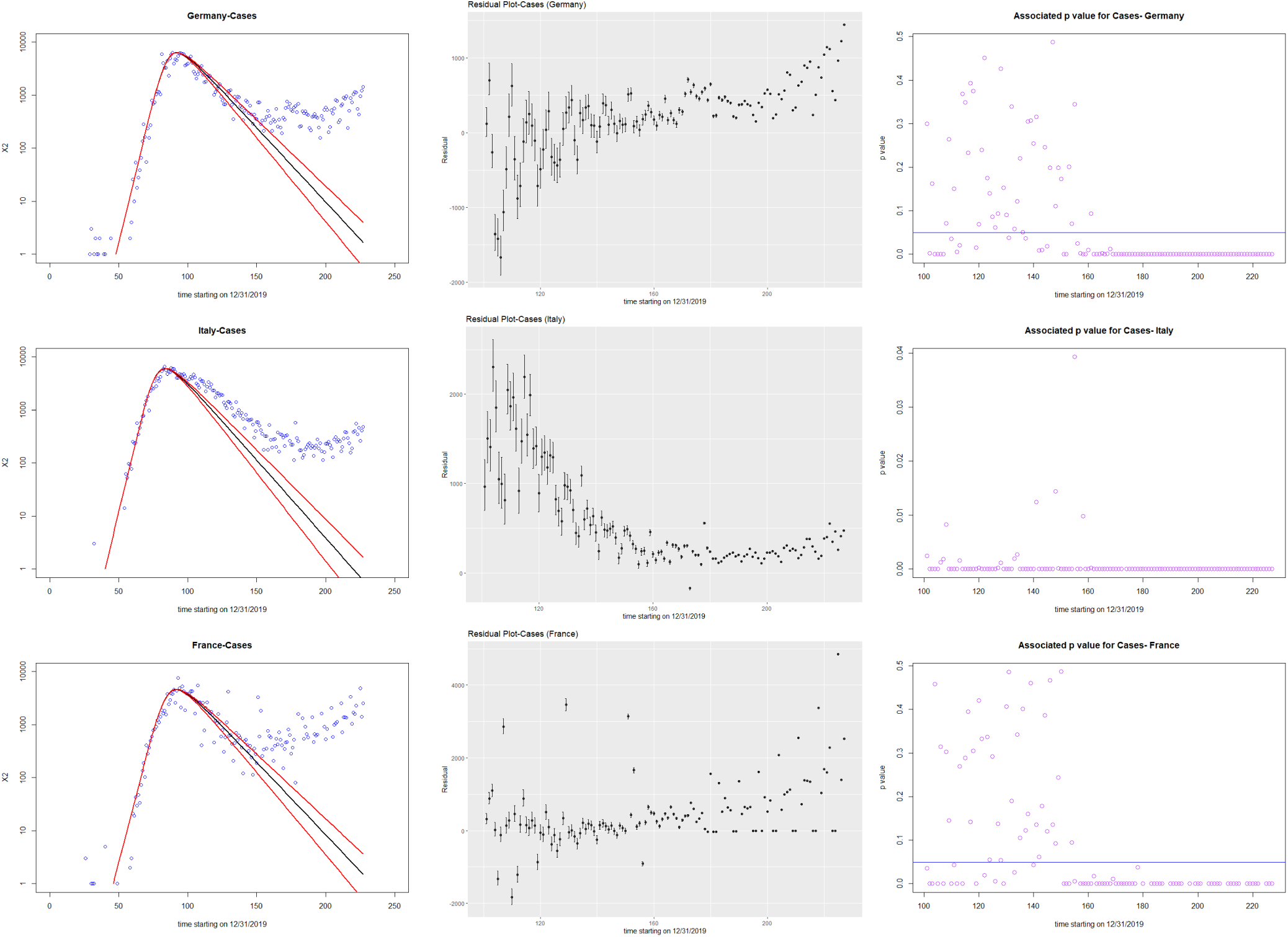
Plots of cases, residuals, and p values against time starting from December 31, 2019. Observed data (blue dots) for the number of cases per day (X2(t)) and fits (solid lines) obtained by solving (3.1) and (3.2) using the ODE solver ode in R. The mean values of the parameters obtained (inset) are from the solid black line and the error bars are from the two red lines. The next plot is the residual plot for the cases, found by subtracting the observed value from the fit value, with an error estimation taken from the values of the fits in the error bars. We can observe the error getting smaller as the expected data from the fits fall below the 100 case mark. The final plot is a measure of the p value for each observed data point given a mean of the middle fit and a standard deviation calculated from the values of the error bars. The horizontal blue line indicates an *α* = 0.05 level of significance, leading us to reject any values below that line as expected. This is replicated for all 9 European countries.

**Figure 2:**
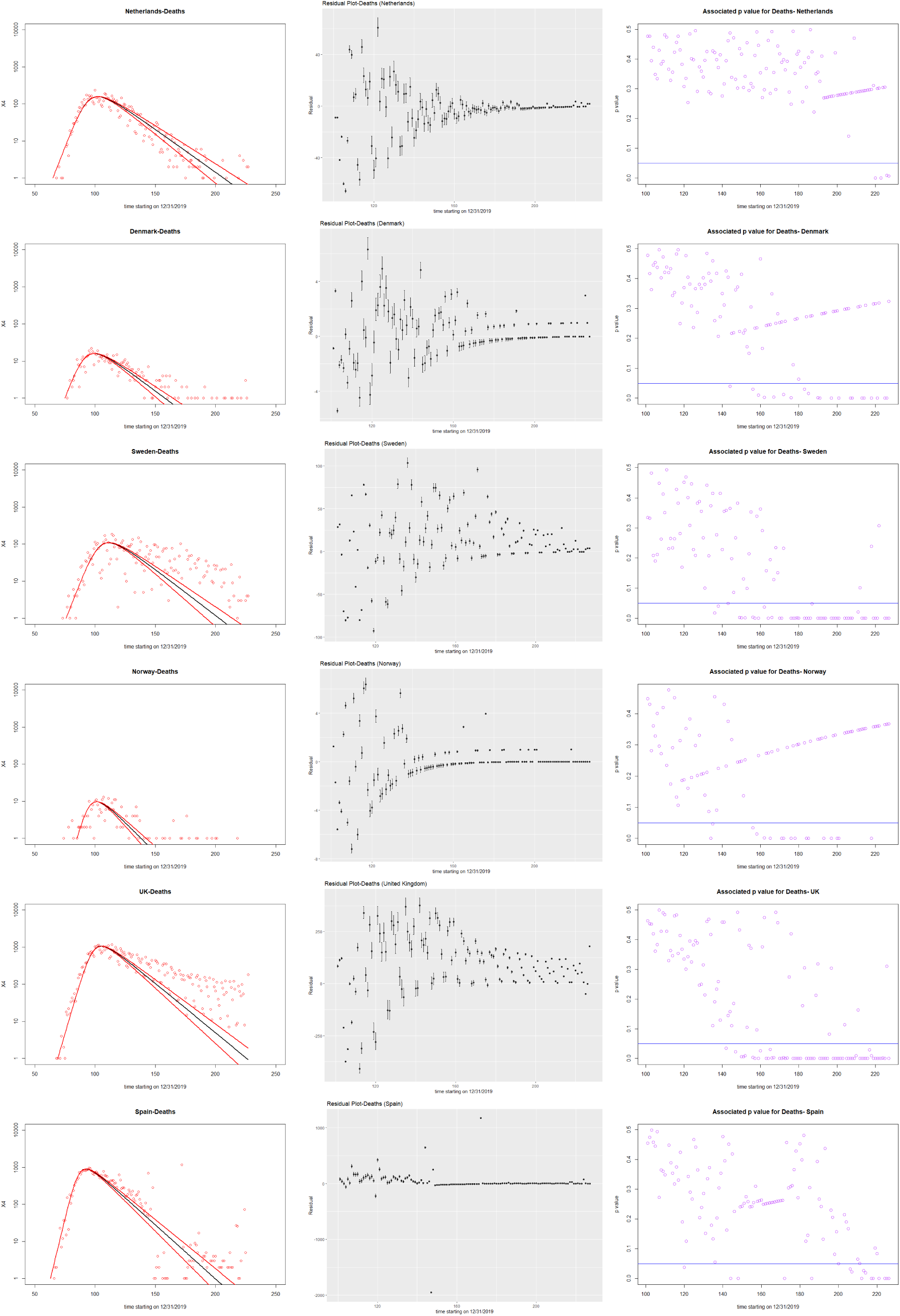
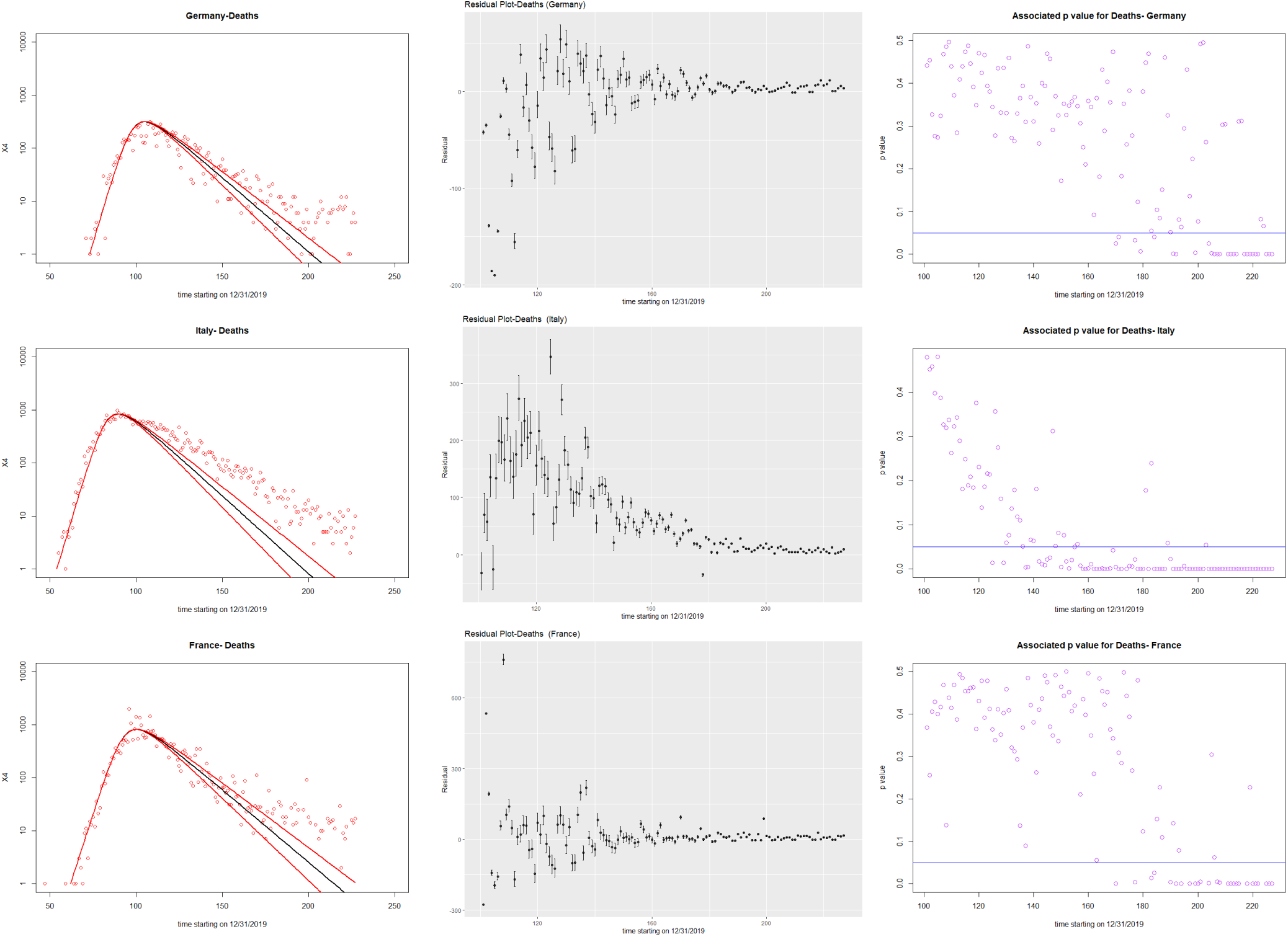
Plots of deaths, residuals, and p values against time starting from December 31, 2019. Observed data (red dots) for the number of deaths per day (X4(t)) and fits (solid lines) obtained by solving (3.1) and (3.2) using the ODE solver ode in R. The mean values of the parameters obtained (inset) are from the solid black line and the error bars are from the two red lines. The next plot is the residual plot for the deaths, found by subtracting the observed value from the fit value, with an error estimation taken from the values of the fits in the error bars. We can observe the error getting smaller as the expected data from the fits fall below the 100 case mark (unless the peak is really small, in which case the error gets smaller as the expected data falls below the 10 case mark). The final plot is a measure of the p value for each observed data point given a mean of the middle fit and a standard deviation calculated from the values of the error bars. The horizontal blue line indicates an *α* = 0.05 level of significance, leading us to reject any values below that line as expected. This is replicated for all 9 European countries.

**Figure 3:**
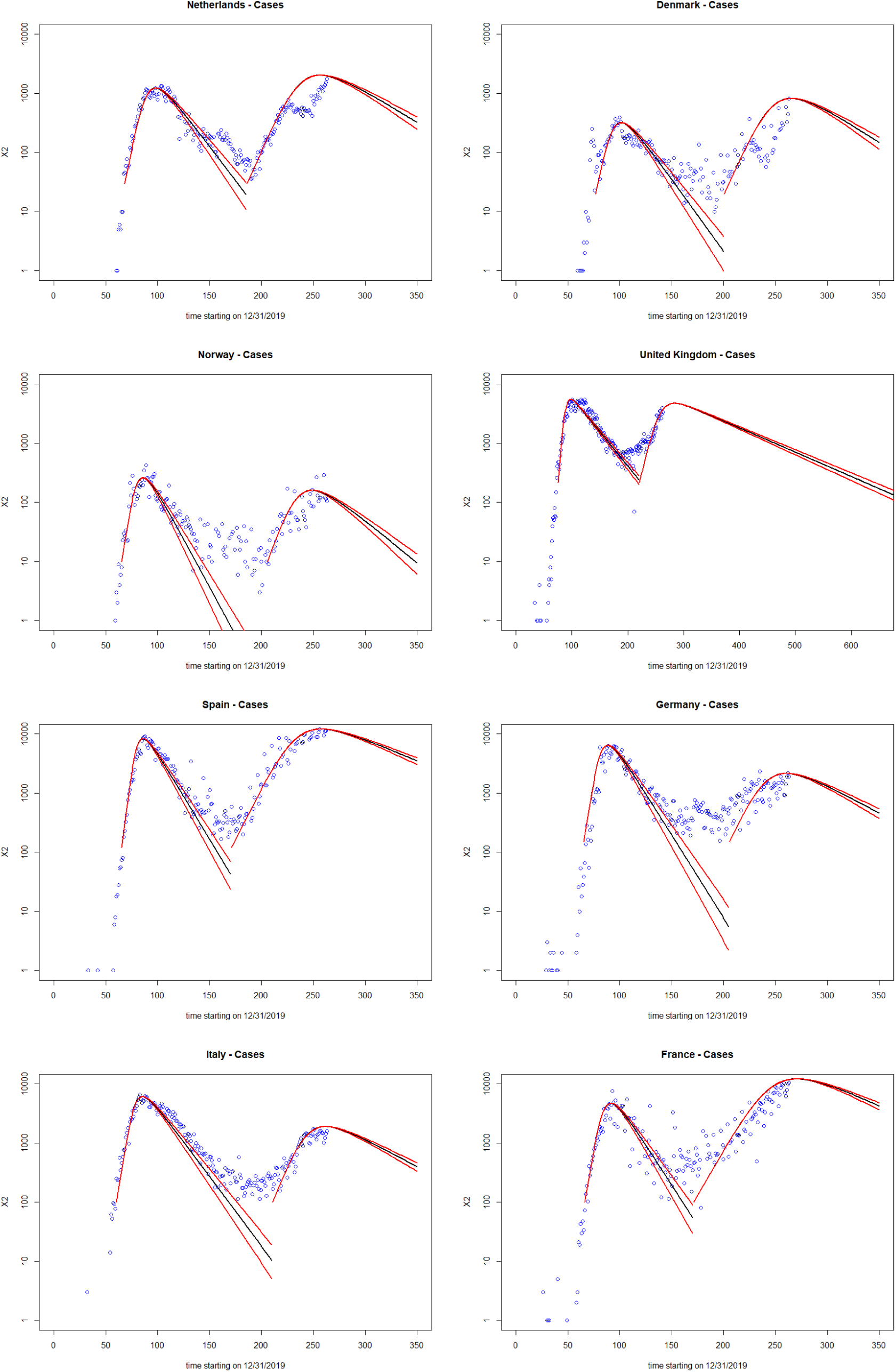
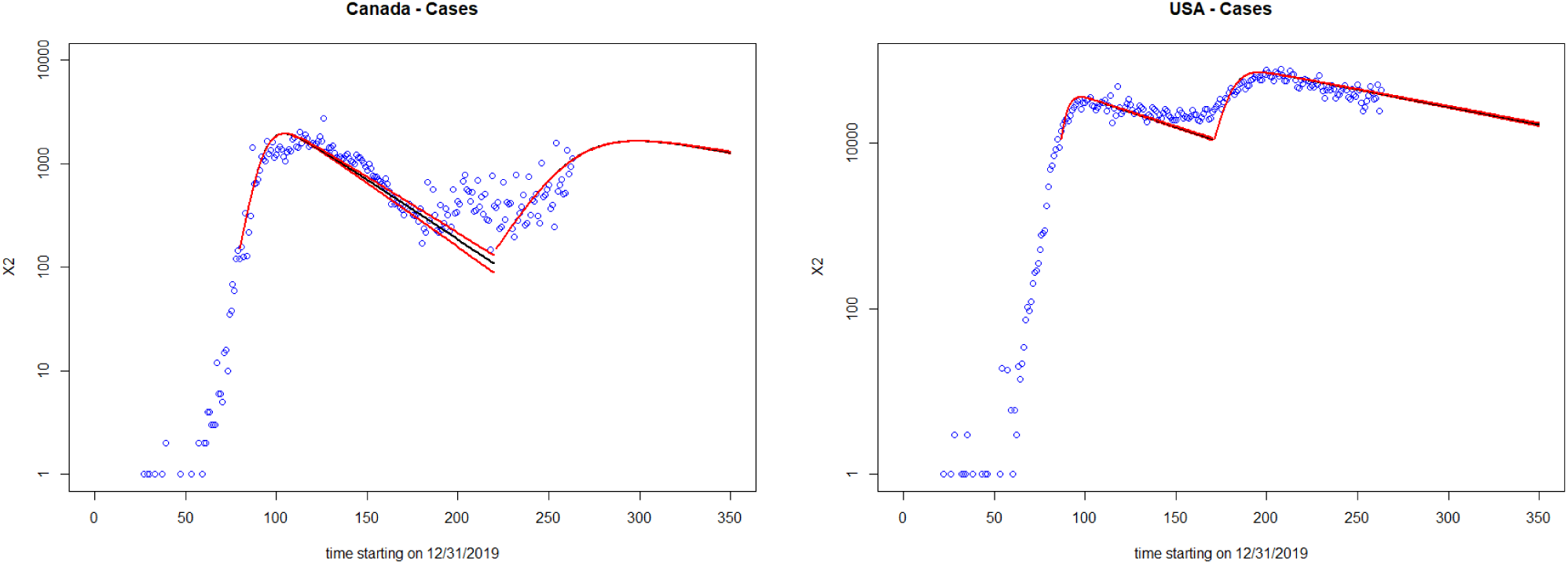
New plot of cases after the lockdown was lifted. Observed data (blue dots) for the number of cases per day (X2(t)) and fits (solid lines) obtained by solving (3.1) and (3.2) using the ODE solver ode in R. The mean values of the parameters obtained (inset) are from the solid black line and the error bars are from the two red lines. This graph contains the model fits for after reopening starts. Data was obtained by ending the first curve at a certain date (found using the residual and p plot analyses) and starting the second curve around that date.

